# Designing a computer-assisted diagnosis system for cardiomegaly detection and radiology report generation

**DOI:** 10.1101/2024.09.02.24311997

**Authors:** Tianhao Zhu, Kexin Xu, Wonchan Son, Kristofer Linton-Reid, Marc Boubnovski-Martell, Matt Grech-Sollars, Antoine D. Lain, Joram M. Posma

## Abstract

Chest X-ray (CXR) is a conventional diagnostic tool for cardiothoracic assessment, boasting a high degree of costeffectiveness and versatility. However, with an increasing number of scans to be evaluated by radiologists, they can suffer from fatigue which might impede diagnostic accuracy and slow down report generation. We describe a prototype computer-assisted diagnosis (CAD) pipeline employing computer vision (CV) and Natural Language Processing (NLP) trained on the publicly available MIMICCXR dataset. We perform image quality assessment, view labelling, segmentation-based cardiomegaly severity classification, and use the output of the severity classification for large language model-based report generation. Four certified radiologists assessed the output accuracy of the CAD pipeline. Across the dataset composed of 377,100 CXR images and 227,827 free-text radiology reports, our system identified 0.18% of cases with mixed-sex mentions, 0.02% of poor quality images (F1=0.81), and 0.28% of wrongly labelled views (accuracy 99.4%), furthermore it assigned views for 4.18% of images which have unlabelled views. For binary cardiomegaly classification, we achieve state-of-the-art performance of 95.2% accuracy. The inter-radiologist agreement on evaluating the report’s semantics and correctness for radiologist-MIMIC is 0.62 (strict agreement) and 0.85 (relaxed agreement) similar to the radiologist-CAD agreement of 0.55 (strict) and 0.93 (relaxed). Our work found and corrected several incorrect or missing metadata annotations for the MIMIC-CXR dataset, and the performance of our CAD system suggests performance on par with human radiologists. Future improvements revolve around improved text generation and the development of CV tools for other diseases.

## Introduction

More than a century has passed since its discovery, still, chest X-ray (CXR) remains a staple non-invasive diagnostic tool in medical institutions across the globe owing to its versatility, inexpensiveness, and convenience (1). Thoracic anatomical structures including the lungs and heart are visualised and distinguished on a 2D film based on radiodensity variations, enabling the detection of common cardiothoracic conditions such as pneumonia, pneumothorax and tuberculosis (2). The procedure costs only a fraction of more sophisticated imaging techniques such as computer tomography (CT) or magnetic resonance imaging (MRI) and can be completed in mere minutes, making it an ideal starting point in thoracic imaging (3).

Despite its popularity, CXR faces its own challenges in its application. Unlike CT and MRI, CXR reduces the 3D pleural space to a singular plane, leading to structural overlaps between anatomical features, with over 40% of the lung parenchyma obscured by the ribs and mediastinum (4). Depending on the type and site of a lesion, the visible differences between the lesion and surrounding structures may be extremely subtle or even indistinguishable (5, 6).

Apart from technical limitations, human factors also complicate CXR interpretation. Radiologists are subject to cognitive biases, distractions, and fatigue, which negatively impact their diagnostic acumen and brood errors, potentially leading to patient harm (7, 8). Furthermore, the ease of performing a CXR has inadvertently led to widespread over-prescription, with the average US radiologist workload increasing by 30% to 70% in under three decades, contributing to significant radiologist burnouts (1, 9, 10). In less privileged regions of the world, severe radiologist shortages are often observed with practising radiologists struggling to meet demand (11). It is therefore perhaps unsurprising that even with numerous technical advancements and error-mitigating strategies developed over the years, studies have repeatedly failed to identify any improvement in radiology interpretive error rates since 1949, which stands at an uncomfortable 3% to 4% of reports, a sizable fraction of which is comprised of CXR (7, 8, 12).

To remedy this chronic deficiency, exploratory efforts have been made to incorporate artificial intelligence (AI) technology into CXR interpretation in recent years (5). The underlying rationale is that, unlike their human counterparts, AI readers are immune to fatigue, distractions, and may suffer less from certain types of cognitive biases based on the training methods, allowing for higher accuracy and consistency (5). Additionally, AI could significantly relieve the burden of radiologists, especially in times of crisis such as the COVID-19 pandemic, reducing the backlog and improving patient outcome (13, 14).

Due to the sheer complexity of the CXR interpretation, most modern approaches employ deep learning methods, such as training convoluted neural network (CNN) models (5, 15, 16). Nevertheless, attempts to implement AI in radiology reading tend to diverge in their philosophy. Some aim to develop AI-augmented diagnostic tools to assist human radiologist interpretation, while others are concerned with creating a system to automate the entire process. For instance, the CAD system CheXGAT (17) helps radiologists detect and discern between fourteen chest conditions, while AI-CenterNet CXR (18) is a fully automated system for detecting and localising eight thoracic conditions. Similarly, some projects may pursue a comprehensive model for wide-spectrum usage such as the ones above, while others narrow their focus to one or few diseases (5, 19). COVID-Net CXR2 and Covid-MANet, CNN models developed for detecting COVID-19 pneumonia, are examples of the latter (13, 14).

The present study produces a focused CAD system for the automatic detection and reporting of cardiomegaly in CXR. Cardiomegaly refers to abnormal or pathological enlargement of the heart, which is often a manifestation of dilated or hypertrophic cardiomyopathies (20–22). While the aetiology of both cardiomyopathies is highly complex and not yet fully elucidated, it is known that both could lead directly to heart failure syndrome which carries a poor prognosis (20, 23, 24). Severe cardiomegaly on CXR is immediately apparent to experienced radiologists on a Posterior-Anterior (PA) CXR, while more ambiguous cases may require the calculation of cardiothoracic ratio (CTR) using a PA CXR (Fig 1). Any value above a threshold (often set between 0.45∼0.55) possibly indicates cardiomegaly (20, 25, 26). The process can be time-consuming and contain a degree of subjectiveness due to variations in thresholds used and measurements taken, and thus may benefit from the implementation of CAD systems.

**Fig. 1.**
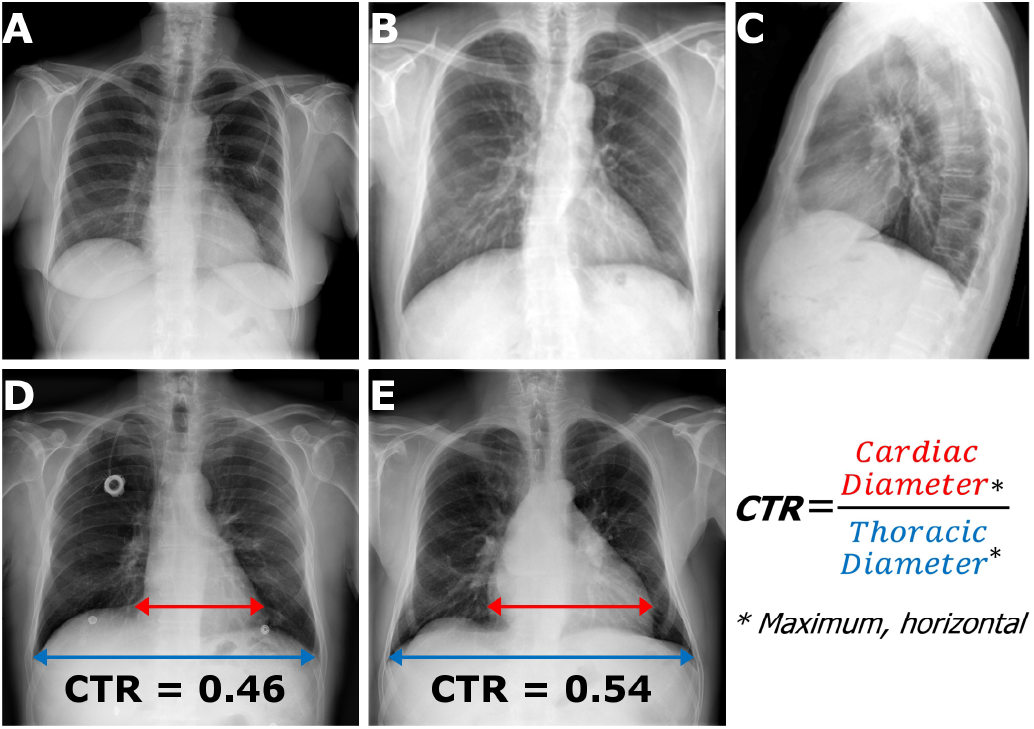
Chest X-ray views and cardiothoracic ratio calculation from frontal images. A: Frontal (Posterior-Anterior, PA) chest X-ray image. B: Frontal (Anterior-Posterior, AP) chest X-ray image. C: Lateral chest X-ray image. D: Frontal chest X-ray image with normal heart size with the two components of the cardiothoracic ratio (CTR) indicated. E: Frontal chest X-ray image with enlarged heart size (CTR ≥ 0.5). A, D, E: images from (34). B, C: images from (35).

Several CAD systems have been developed for cardiomegaly detection and can be categorised into classification-based and segmentation-based approaches (27). Several transfer learning models were developed, with one classifying normal vs cardiomegaly CXR images with an AUC of 0.87, which is close to the average of similar approaches (28– 30). U-Net-based segmentation models are utilised to automatically calculate CTR for determining cardiomegaly presence and are considered a feasible option with the advantage of higher explainability over classification-based methods (31, 32). Nevertheless, the existing cardiomegaly CAD systems share many of the same limitations. The datasets used for model creation and testing are often limited in size due to the rarity of pixel-level labelled CXR data, restricting model generalisability (27, 32). The final output of these models is either directly binary, or CTR ratios which are then used to derive binary decisions (presence or absence of cardiomegaly) (27, 33). Without the capability of determining the severity of heart enlargement or producing report-styled sentences mimicking those written by human radiologists, the capacity to which these CAD systems can automate cardiomegaly detection and reporting is limited.

The current study is inspired by existing segmentation-based cardiomegaly CAD systems and improves upon them by enhancing generalisability and introducing missing features crucial to realising full automation in cardiomegaly detection and reporting. MIMIC-CXR, a massive dataset containing 377,100 CXR images and 227,827 reports, is selected as the main dataset. A ResNet50 model is first trained using the dataset for filtering CXR images suitable for cardiomegaly detection. Lung and heart segmentations and CTR calculation are subsequently performed on suitable images with varying degrees of cardiomegaly reported. Statistical techniques are then applied to determine the CTR threshold for different severity levels of cardiomegaly, ranging from borderline to severe. Finally, natural language processing (NLP) and large language models (LLM) are leveraged to produce accurate, believable cardiomegaly report sentences based on machine predictions. If fully ensembled, the CAD system only requires an input stream of CXR images and provides full automation from image quality assessment to report sentence generation, subjective to double checking by human radiologists, as illustrated by the flowchart (Fig2). A cohort of four radiologists were invited to assess the output quality of CAD output sentences as human experts.

**Fig. 2.**
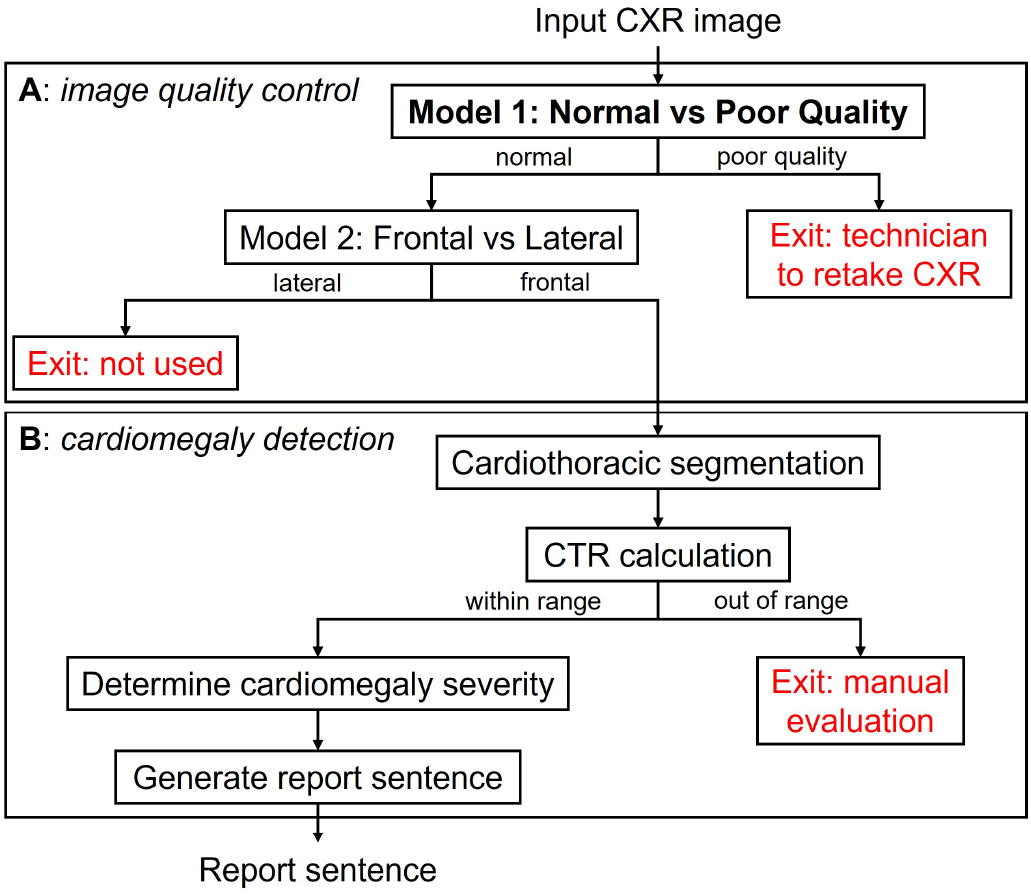
Flow chart of the computer-assisted diagnosis pipeline. A: Image classification models. B: Image segmentation and sentence generation models. Indicated in red are ‘exit’ steps where the image or result cannot be used by the pipeline.

We hypothesise that coupling the output of computer vision models with large language models can generate plausible expert-level radiology report text. Specifically, we aim to:

1) assess the quality of the MIMIC-CXR dataset, including both images and reports, to prepare it for developing computer vision models, 2) Utilise CXR segmentation algorithms to derive features relevant to sub-tasks, 3) create a computer vision model for cardiomegaly classification using labels extracted from reports using natural language processing tools, 4) combine the output of the computer vision classification model with large language models to generate plausible medical text, and finally 5) use expert evaluation of the generated reports to assess the quality of the generated text.

## Materials and Methods

### MIMIC-CXR dataset

The MIMIC-CXR database (36) was selected as the primary source of CXR data in this study. It is publicly available with credentialed access via PhysioNet (37) and includes 377,100 CXR images in both JPG and DICOM format and 227,827 corresponding free-text radiology reports, all anonymised. The images and corresponding text reports are stored across 10 folders (p10-p19). Additionally, the database contains CSV files with image metadata and feature annotations acquired with CheXpert (38) and NegBio (39). In each feature column, 1 represents a positive mention of the feature, 0 represents negative mentions, and −1 represents ambiguous mentions. A blank value indicates the absence of any mention of the feature.

All CXR images in JPG format and their corresponding free text reports, alongside metadata and annotation CSV files, were directly downloaded from PhysioNet using command prompts.

The data user agreement of MIMIC-CXR prohibits sharing images from the dataset. Images from MIMIC-CXR that are already in the public domain also fall under this agreement. The MIMIC-CXR team have confirmed that this policy applies to all data therefore no disclosure is permitted (private communication 30 August 2024). We therefore have replaced the images with similar images from the NIH Chest X-ray dataset (34) (for frontal PA images) and from (35) (for AP and lateral images, under a CC BY-NC 3.0 license). In each of the figures (where relevant) we have indicated the original MIMIC ID that the inference is made on so that those with credentialised access to MIMIC-CXR can inspect the original data. The report snippets shown here are all synthetically generated, however their context mimics the original MIMIC-CXR report text. MIMIC-CXR report IDs are disclosed in the relevant captions to allow readers with MIMICCXR data access to inspect the original images and reports.

### Generation of poor quality images

All images from p10 and p11 folders were scrutinised manually to identify ‘abnormal’ images which are those that cannot be properly interpreted by radiologists and would warrant retaking the image. These images contain severe cropping, distortion, and/or absence of thoracic structures. Upon positive identification, these images were then used to generate 500 additional ‘artificial’ abnormal images by applying synthetic and augmentation techniques to imitate common types of abnormal images from normal images (see examples in Fig 3A). The ‘genuine’ abnormal images were set aside for testing (see below).

**Fig. 3.**
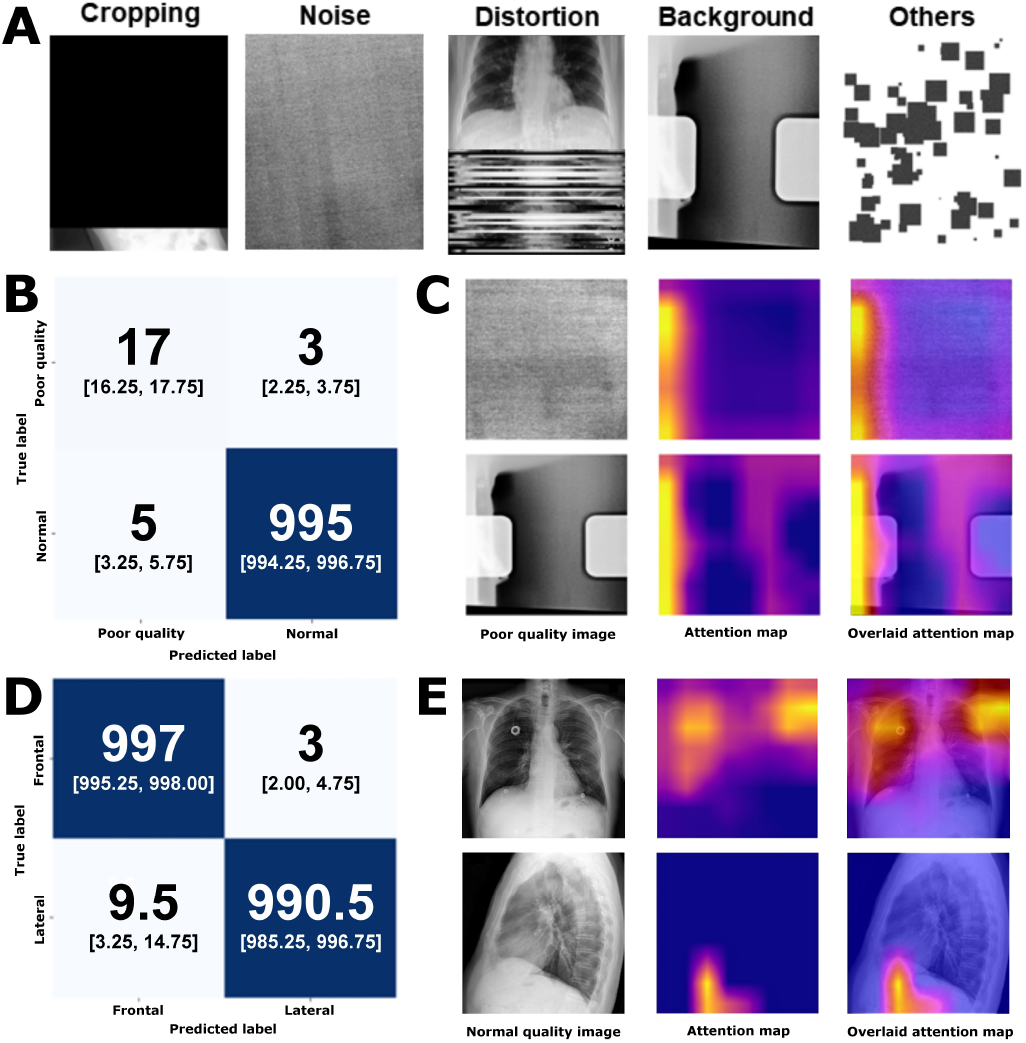
Data description, computer vision model classification results, and model interpretability. A: Generated poor-quality images due to cropping, noise, distortion, background and other types (patching) of image manipulations. B: Confusion matrix of normal versus poor quality classification model with the median [interquartile range (IQR)] displayed across 10 models initialised with different random states. C: Example of poor-quality input images and the model attention map. D: Confusion matrix of frontal versus lateral classification model (median [IQR] across 10 models initialised with different random states). E: Example of frontal (from (34)) and lateral (from (35)) input images and the model attention map.

### Computer vision models

Two classification models were trained on the MIMIC-CXR dataset, one to classify normal from poor quality images, and a subsequent model to classify frontal from lateral images as shown in Fig 2A to perform an image quality control.

### Classification

For the normal versus poor quality image classification model, 500 frontal including both PA and AP views as well as 500 lateral images were selected at random for the normal group from the p10 and p11 folders to form the training set, where each image was manually checked.

For the frontal versus lateral classification model, 1,000 images from each group were selected from the entire MIMICCXR dataset (p10-p19) through cross-referencing with view labels reported in the metadata CSV file. Similarly, each image was manually checked for labels being correctly assigned. The test set consists of another 2,000 images (equally split) selected at random, and manually checked, without overlapping patients with the training set.

For both models, the training image datasets were randomly split into training and validation sets at a ratio of 80:20, enabling real-time model training performance monitoring. The ResNet50 model (40) was loaded without pre-trained weights, and all images were resized to 256×256 pixels. Data augmentation was incorporated into training data loading which randomly introduced contrast jitter and rotations to all images for boosting data variability, model versatility and robustness. Multiple combinations of training parameters and hyper-parameters were experimented with for performance optimisation in a grid search-like process with manual tuning. For both training and validation sets, the batch size was set to 64, with cross-entropy loss and Adam optimiser (learning rate of 0.001). 20 epochs were scheduled, with an early stopping condition if both training and validation accuracy surpassed 99% at the end of an epoch. The trained model states were saved as .pth (PyTorch-trained model) files. Each model was initialised 10 times with different random states and results are represented as median with interquartile range.

The trained models were subsequently evaluated on the independent test sets. Performance was gauged by overall accuracy, as well as precision and recall, with confusion matrices constructed. To understand the rationale of the model in classifying images, Grad-CAM (41) was used to create attention heatmaps to visualise the focus areas of each model, providing insight into what features it considers distinctive to each class, as well as catching instances where the model may predict based on non-relevant features.

### Segmentation and cardiothoracic ratio calculation

Good quality frontal CXR images with cardiomegaly mentions in their associated reports were then grouped based on the severity descriptors extracted from text reports. For each frontal image, segmentation of the lungs and heart was carried out using the inbuilt default U-Net-based segmentation model in the Chest X-ray Anatomy `Segmentation` (CXAS) package (42) (v0.0.9), generating lungs and heart masks in the PNG format.

The Maximal Horizontal Diameter (MHD) of the lungs and heart masks were used to calculate the computer vision-based CTR (CV-CTR). The MHD was calculated by counting the number of mask pixels in each row of a given mask image using `opencv-python` (v4.9.0.80) and `numpy` (v1.26.2) packages, and subsequently taking the 95^th^ percentile value to eliminate potential outliers caused by sub-optimal segmentation.

If either the lung or heart MHD of an image was 0, it was excluded from further analysis as segmentation had likely failed for that image. The CV-CTR for each image was then calculated by dividing the heart MHD by the lung MHD (Fig 1). The CV-CTRs were subsequently grouped by severity descriptors from the reports.

Additionally, another set of ratios was calculated using the total areas, instead of HMD, of heart and lung masks. Areas were measured by counting the total number of mask pixels within lung and heart masks, and the Cardiothoracic Area Ratio (CAR) was calculated by dividing heart by lung area.

### Natural language processing tools

#### Extracting data from reports

CXR reports containing either positive or negative mentions of cardiomegaly and its synonyms were filtered by selecting only reports labelled 1 or 0 in the ‘cardiomegaly’ column in the CheXpert CSV, thereby rejecting any reports with ambiguous mentions or no mentions concerning cardiac size. For each selected report, NLP methods, including text tokenisation and keyword searching, were applied using the NLTK (v3.8.1) package to extract sentence(s) describing cardiomegaly or cardiac size. Cardiomegaly mentions were extracted from a total of 137,424 reports through this process, roughly 60% of all MIMICCXR dataset reports.

All adjectives and adverbs from extracted cardiomegaly mentions were identified and ranked based on frequency. From the top 50 most frequent adjectives and adverbs respectively, severity descriptors (terms that describe the severity of a condition) that appeared in at least 100 reports were selected for further analysis. Adjectives and adverbs sharing the same stem (such as mild and mildly) were merged. Cardiomegaly mentions files containing these severity descriptors were grouped by keyword matching, with any files containing two or more different severity descriptors being discarded.

#### Comorbidity analysis

Comorbidity analysis was performed for each report using annotations of 14 conditions in the CheXpert CSV. The conditions were ‘Atelectasis’, ‘Cardiomegaly’, ‘Consolidation’, ‘Edema’, ‘Enlarged Cardiomediastinum’, ‘Fracture’, ‘Lung Lesion’, ‘Lung Opacity’, ‘No Finding’, ‘Pleural Effusion’, ‘Pleural Other’, ‘Pneumonia’, ‘Pneumothorax’, and ‘Support Devices’. Any positive mention of the condition in the report (labelled ‘1’ in the CSV) was considered an occurrence. Ochiai similarity (Eqn. 1) was calculated between conditions to analyse their degree of association (where *a* is the number of co-occurrences of conditions A and B, *b* the number of single occurrence of condition A, and *c* the number of single occurrence of condition B). We used Ochiai similarity instead of co-occurrence frequency as different conditions had varying occurrence frequencies. The Ochiai similarities among the 14 conditions were used to construct a graph of co-occurrences using the `NetworkX` package (v3.1).

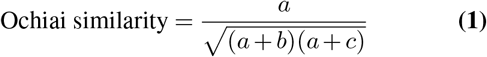

#### Report text generation

Previously extracted cardiomegaly mentions were regrouped with severity categories. The mentions were pre-processed to strip periods and all letters were converted to lowercase to account for minor variations in expressions. Within each severity category, the most frequently used sentences for cardiomegaly reporting were counted and ranked. Expressions that mention other conditions irrelevant to cardiomegaly and/or patient history were filtered out. The ten most frequent filtered expressions from each category were given to MedQBot on Poe (https://poe.com/medqbot), an LLM originally intended for medical exam preparation, with a prompt to generate ten similarly concise, report-styled, cardiomegaly expressions. The prompt used is listed below, where [severity group] was replaced by each of the adjectives or adverbs identified above:

> *Here are some CXR report sentences describing* [severity group] *cardiomegaly. Based on this set of sentences, generate similarly concise and report-styled sentences describing* [severity group] *sentences*.

The generated sentences were acquired on 13 March 2024. They were manually checked for accuracy, style and relevance, with unsuitable sentences (e.g. too verbose, unrelated, inaccurate, temporal) being discarded.

#### Human evaluation

To evaluate our cardiomegaly CAD pipeline performance, four radiologists (three with 1 to 5 years of experience, and one with 10+ years of experience) were invited to participate in a small-scale assessment using the online Qualtrics questionnaire. A total of 40 items were prepared, each consisting of a frontal CXR image, a report snippet of the findings section, and a side-by-side question box where the radiologists indicate to what degree they agree with each sentence from the snippet based on the image provided. The radiologists were able to provide reasoning for any disagreement in a free-text box. Each item was seen by two radiologists who were blinded to each other’s answers and were not informed whether the sentences were machine-generated or from the original MIMIC report. Each radiologist evaluated a similar number of sentences, with the same ratio of changed and unchanged sentences.

The questions were divided into four categories (Table 1) to ensure an even mix of original and machine-generated sentences, either related or unrelated to cardiomegaly. The machine-generated cardiomegaly sentences were randomly selected from the pre-curated list as described before based on the predicted severity group by the computer vision model. For question categories 2 and 4, non-cardiomegaly sentences were rewritten and paraphrased with MedQBot with the following prompt on 19 March 2024:

> *For the following paragraph taken from a CXR report, generate a new paragraph by paraphrasing each sentence from the given paragraph, while maintaining concision and standard CXR reporting style*.

**Table 1.** Question composition of radiologist questionnaire. Where ‘unchanged’ indicates the sentences are those from the original MIMIC-CXR reports, ‘CAD system’ indicates cardiomegaly sentences are generated with the pipeline described above, ‘MedQBot’ indicates sentences generated by LLM-paraphrasing.

| Category | n | CM sentences | Other sentences |
| --- | --- | --- | --- |
| 1 | 8 | Unchanged | Unchanged |
| 2 | 8 | Unchanged | MedQBot |
| 3 | 12 | CAD system | Unchanged |
| 4 | 12 | CAD system | MedQBot |

Each radiologist was assigned 20 items (each containing multiple statements to assess), with each radiologist receiving roughly the same number of items in each listed category. The assignment process also ensured that each question was answered by exactly two radiologists. The radiologists were not informed of the design or pipeline of this study nor the composition of the question set or that of their assigned questions. Their responses were individually downloaded as CSV from Qualtrics and pre-processed to integrate into a single data frame using pandas (v2.1.4) with paired responses for each question, complete with question category and cardiomegaly relevance.

To assess inter-rater agreement we computed the total agreement by comparing their results for each sentence. We used two agreement metrics: strict agreement which consisted of four distinct categories (‘Agree’, ‘Partially Agree’, ‘Disagree’, and ‘Unsure’) and relaxed agreement which merged ‘Agree’ and ‘Partially Agree’ into a single category.

## Results

### Data quality assessment

We first reviewed the data quality for several aspects relevant to this study. The MIMICCXR dataset is one of the largest publicly available chest X-ray radiology datasets, thus containing a considerable degree of data variability. Consequently, only a subset of the MIMIC-CXR dataset is suitable for developing cardiomegaly detection CAD systems.

Dataset filtering for downstream applications is complicated by a host of factors. As mentioned before, frontal CXR is best suited for cardiomegaly detection, while lateral images are rarely used and should thus be excluded. Furthermore, the MIMIC-CXR dataset is not error-proof. We found that 421 patient reports contain mismatched sex mentions (Table 2). View labels of images were occasionally missing or incorrect, for instance, by mislabelling a lateral image as PA/AP. In addition, a smaller proportion of images suffer from quality issues severe enough to entirely prevent analysis. To address this, the CAD system needs to first identify and flag unsuitable images to prevent further processing and analysis while alerting human radiologists. This is accomplished by leveraging two ResNet50 classification models as detailed below.

**Table 2.** Data quality assessment of the MIMIC-CXR dataset (65,379 patients, 227,827 individual reports, 377,100 images). Indication of mismatched sex mentions in reports attributed to the same individual, number (%) of poor quality images indicated by our poor quality image classification model, and number (%) of wrongly labelled views (in the metadata) indicated by our view classification model. All reports and images indicated above were manually checked, and we provide a spreadsheet in the Supplementary Information with the corrected view labels and reports likely from different individuals due to sex differences with other reports attributed to the same person identifier.

| Data quality assessment category | n | % |
| --- | --- | --- |
| Patient reports with mixed sex mentions | 421 | 0.18 |
| Poor quality images | 83 | 0.02 |
| Wrongly labelled views | 1,054 | 0.28 |
| Unlabelled views | 15,769 | 4.18 |

### ResNet50 classification sub-models and metadata quality

As mentioned, a two-step hierarchical model was developed to classify CXR images by quality and view for dual purposes. In direct relation to the CAD system itself, the model provides a filtering step to only allow suitable images to be assessed by downstream cardiomegaly detection algorithms. Additionally, the model can also be utilised to help correct mislabelling and flag flawed images within the MIMIC-CXR dataset, or potentially any similar CXR datasets. 22 abnormal images were identified from p10 and p11 subfolders and categorised into five groups based on similar features (Fig 3A). For each category, 100 new images were generated with synthetic and augmentation techniques. Together, they constituted an abnormal training subset of 500 images to train the first model to classify normal vs abnormal images. This model demonstrated high overall accuracy on a test set with 20 real bad-quality images from the MIMIC-CXR and 1,000 normal images with 85% recall for poor quality class (Fig 3B). The second model, which differentiates between frontal and lateral CXR, exhibited much better performance, with 99.4% accuracy on the test set with 1,000 frontal and lateral images each (Fig 3D).

For both sub-models (see Fig 2), model explainability was explored through mapping model attention with Grad-CAM (Fig 3C, E). Model attention for sub-model 1 is relatively localised with a region of high intensity on the left. For submodel 2, model attention is diffused when frontal images are encountered, covering most of the image with higher attention zones covering both lungs. When presented with a lateral image, model attention is markedly more focused and often localised to the region just below the diaphragm.

### Cardiomegaly severity classification

#### Extraction of cardiomegaly mentions using natural language processing

Gaining a comprehensive understanding of the cardiomegaly landscape in the MIMIC-CXR dataset is paramount to CAD development. ‘Normal’ is the most common descriptor for heart size with 10,883 cases, followed by ‘Mild’ (4,651 cases), ‘Borderline’ (4,172 cases) and ‘Moderate’ (2,392 cases) when describing positive cardiomegaly. Generally, descriptor frequency decreases with increasing perceived severity. ‘Mild to moderate’, ‘Moderate to severe’ and ‘Severe’, are less commonly used, each with less than a thousand cases (Figure 4A).

**Fig. 4.**
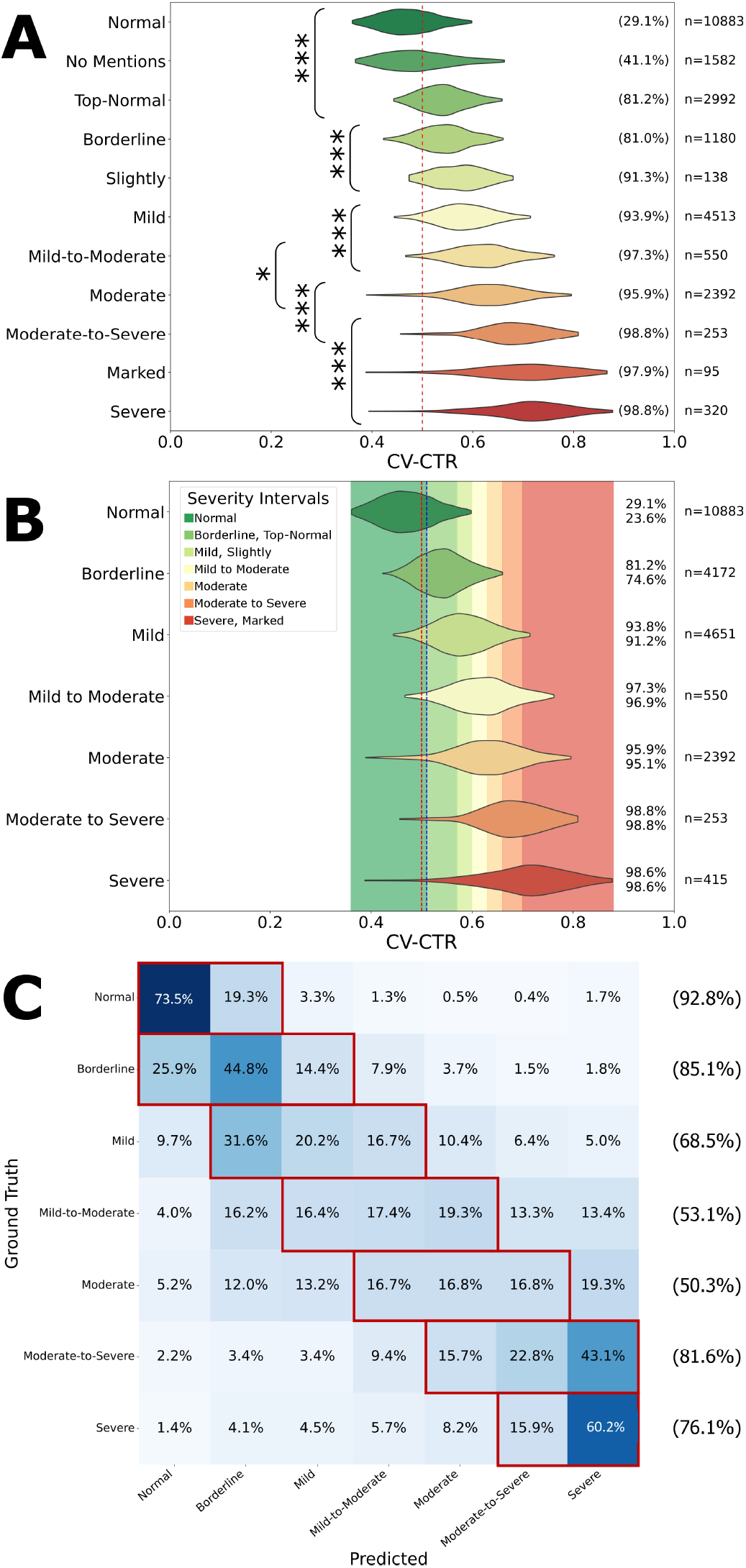
Data-driven severity classification based on natural language extracted labels and computer vision-based cardiothoracic ratio calculation. A: Violin plot of the cardiothoracic ratio (CTR) based on the computer vision (CV) segmentation model. Each adjective/adverb relating to cardiomegaly was normalised (i.e. ‘mild*ly* ‘ to ‘mild’). Also showing reports without cardiomegaly mentions. Comparison of CV-CTR across different severity indicators. * = P<0.05, *** = P<0.001. The percentage on the right-hand side indicates the proportion of images with a CTR over 0.5 (red dashed line). B: Cardiomegaly severity classification model with thresholds determined by the model, grouping of severity indicators with non-significant different CV-CTRs between them. The blue line indicates the cut-off for which the probability of borderline cardiomegaly is larger than normal heart size. The top percentage on the right-hand side indicates the proportion of images with a CTR over 0.5 (red dashed line), whereas the bottom percentage indicates the proportion of images with a CTR over 0.51, the threshold between normal and borderline heart size (blue dashed line). C: Confusion matrix of the CV-CTR classification model into the severity classes. Red boxes and percentages indicate the total number predicted within one category of the labels from radiology reports.

#### CXR auto-segmentation and cardio-thoracic ratio calculation

After CV-CTR calculation, severity descriptors were ranked by median CV-CTR and CAR. For each pair of neighbouring descriptors, t-tests were performed with Šidák correction to identify statistically synonymous severity descriptors which were then merged to form severity categories.

Threshold values separating each severity level were calculated separately for both CV-CTR and CAR. For the former, t-tests were performed for every value between 0 and 1 at intervals of 0.01 with CVCTR data of each severity group, and each value was assigned to the severity group with the greatest p-value, forming ratio intervals. For CAR, t-tests are performed in a similar manner with the range of tested values set between 0 to 0.5 instead. A correlation test between the two sets of ratios was also carried out to determine whether the ratios are correlated on a case-by-case basis.

For each descriptor, CV-CTR was calculated for all corresponding PA CXR images with the computer vision model (Fig 4A). Distributions appear to be normally distributed, although some are slightly skewed, such as the ‘moderate-to-severe’ category. Data spread is generally bigger in higher severity groups. The ranking of descriptors by median is mostly in line with linguistic expectations. 93.8% of mild cardiomegaly cases were found to have CV-CTR over the clinical threshold of 0.5 based on the computer vision model, with upwards to over 98% in severe cardiomegaly, indicating high recall of the CAD system for cardiomegaly cases.

#### Data-driven model for severity classification

Significant overlaps are observed between the distribution of neighbouring descriptors. To identify statistically synonymous severity descriptors, multiple t-tests were carried out between CTR distributions of each pair of neighbouring descriptors, and any neighbouring descriptors with non-significant differences were merged into groups, forming a total of seven severity categories (Fig 4A).

CV-CTR intervals for each severity group were subsequently established for each severity category (Fig 4B). The upper threshold for ‘normal’ CTR (0.51) is close to 0.5, mirroring the clinical threshold for cardiomegaly diagnosis. The ratio intervals for normal and severe categories are much larger than intermediates to encompass extreme values. The number of cases falling into each severity interval varies slightly compared to the reference distribution of cases based on original report descriptors.

Using previously established severity group CV-CTR intervals, a confusion matrix was constructed to evaluate heart size predictions of the model against MIMIC ground truths (Fig 4C). At both ends of the severity spectrum, the model performs relatively well in accurately determining cardiomegaly severity. Intermediate severity categories, such as ‘moderate’, saw a drop in accuracy in severity classification, although the model is still highly accurate for binary classification (normal vs cardiomegaly) across all categories excluding ‘borderline’.

The severity classification based on CV-CTR is similar to the cardiothoracic area ratio (CAR), defined here as the ratio of the total heart area and lung area as presented in PA CXR images. We found the distribution pattern of CV-CAR to be highly similar to that of CV-CTR, and the two are found to be positively linearly correlated (R^2^=0.82), with 90% of residuals within *±*0.09, and with the CV-CAR showing similar patterns across the 7 severities (Supplementary Materials, Figure 7).

#### Report sentence generation using a medical large language model

Once a severity category prediction has been made by the computer vision model, it is used as an input argument for report sentence generation using LLMs. A sentence is picked at random from the pre-curated list of generated sentences of the corresponding severity category to constitute the final report sentence. Fig 5 illustrates three examples of report sentence generation based on cardiomegaly severity predictions. Model prediction for the CXR image in Fig 5A matches that of the original report, and as such a sentence resembling the original is generated. Model prediction for the CXR image in Fig 5B differs from the original by one level of severity and thus is regarded as a close match, while a mismatch is found for the CXR image in Fig 5C.

**Fig. 5.**
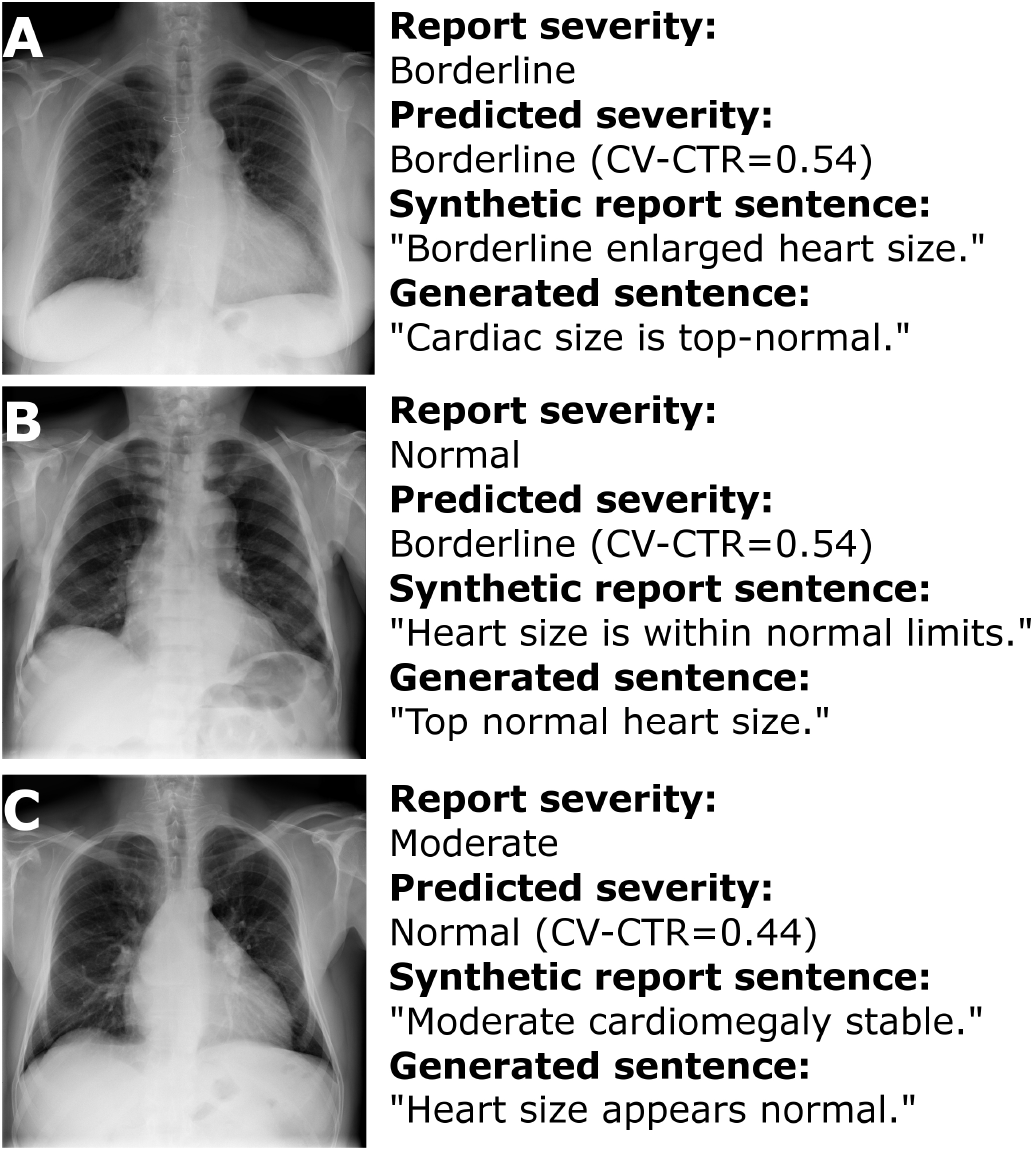
Examples of computer vision-based cardiothoracic ratio calculation and subsequent report text generation. A: Exact match (image from (34); see MIMIC report s56175428 for original report sentence and associated image). B: Close match (within 1 category of report text) (image from (35); original MIMIC report s52317659). C: Mismatch between CAD system and image (image from (34); original MIMIC report s57481090).

### Human evaluation of radiology reports

#### Inter-rater agreement

We calculated the strict and relaxed agreement in three different settings: (1) between our radiologists (strict: 0.487, relaxed: 0.864), (2) between our radiologists and the original MIMIC radiologists on unchanged sentences (strict: 0.624, relaxed: 0.845), and (3) between our radiologists and machine-generated sentences (strict: 0.545, relaxed: 0.929).

Notably, the machine-generated sentences achieved a strict agreement of 0.545, which was 0.089 lower than the maximum strict agreement between our radiologists and the MIMIC radiologists (0.634). In contrast, the machine-generated sentences achieved the highest relaxed agreement (0.929), surpassing the agreement between our radiologists and the MIMIC radiologists by 0.084.

#### Comorbidity mentions

We evaluated the relationship between other mentions in the reports and cardiomegaly. We show here the most common co-occurrences across all reports, and for those with co-occurrences with cardiomegaly (Fig 6A) we visualise the CV-CTR in Fig 6B. Between 53% (lung lesion) and 90% (edema) of patients had these as comorbidities with presence of cardiomegaly.

**Fig. 6.**
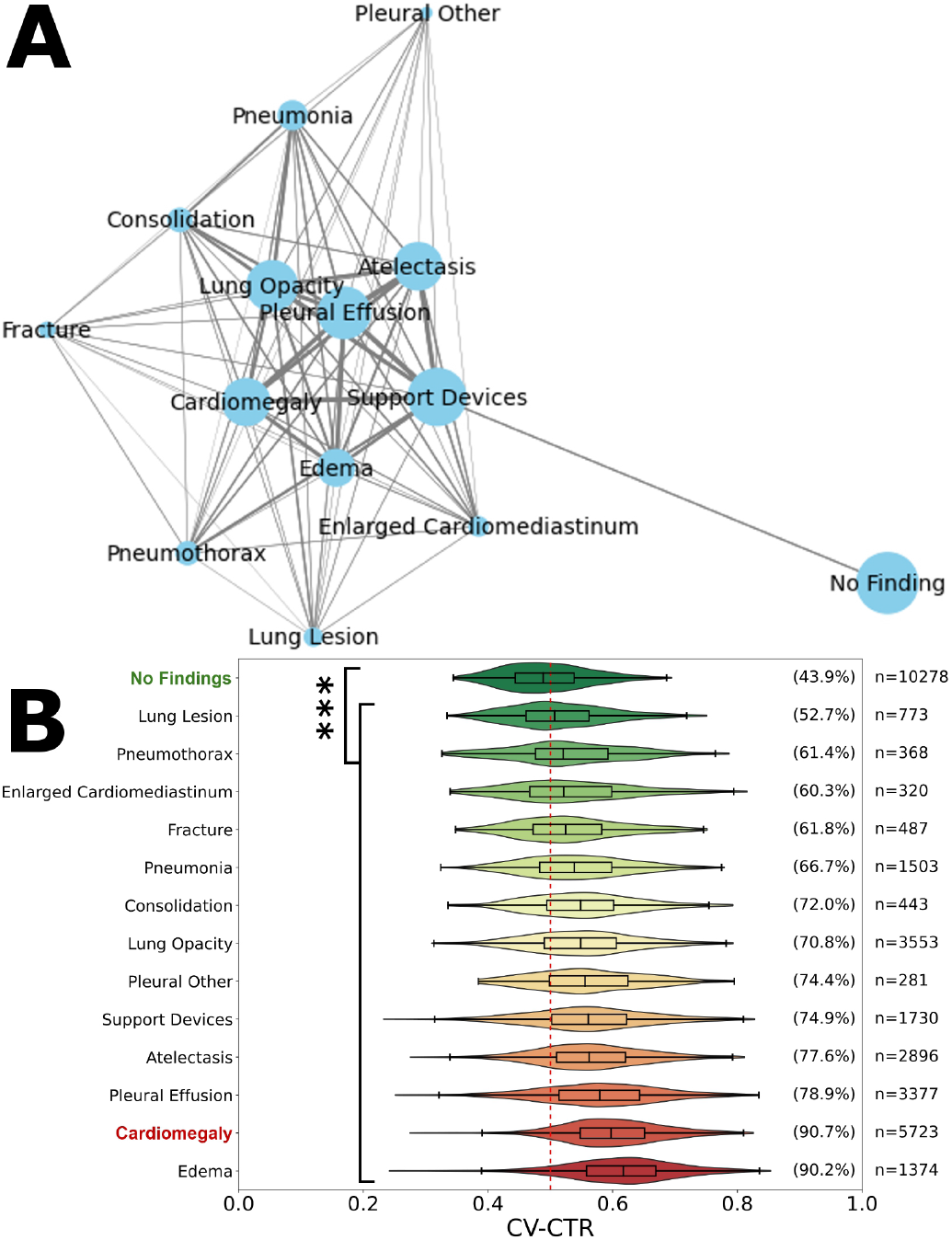
Evaluation of the cardiothoracic ratio and comorbidities with cardiomegaly. A: Co-occurrence graph of the other 14 CheXpert observations by report, including cardiomegaly. Node size indicates the number of reports where the condition is observed. Line width indicates the association between conditions measured by Ochiai similarity. B: Violin plot of computer vision-based cardiothoracic ratio for the most common co-occurrences with other mentions. The percentage on the righthand side indicates the proportion of images with a CTR over 0.5 (red dashed line). Significant differences from the reference group noted, *** = P<0.001.

## Discussion

### MIMIC-CXR Dataset Quality Control

The presence of quality issues, such as abnormal images and mislabelling within the MIMIC-CXR dataset, is an unexpected finding despite the relative rarity. Regardless of the underlying causes, if left unchecked and unbeknownst to researchers, they could pose significant challenges to ensuring data quality and reliability in future studies utilising the dataset.

Current literature involving MIMIC-CXR, however, rarely mentions the dataset quality shortcomings, an odd predicament given the popularity of the dataset. The original journal describing the MIMIC-CXR dataset does contain a brief notice of the variability of images caused by image rotation, poor patient positioning and secondary collimation, yet did not comment on the presence of severe image quality issues nor instances of mislabelling (36). Out of five studies published over the past two years involving the use of MIMIC-CXR images for creating deep learning models, none has acknowledged any aspects of quality issues within the dataset (43–47).

The current study sought to ascertain the extent of selected quality issues within MIMIC-CXR and rectify them for the benefit of future studies using the same dataset. Manual identification of bad-quality images or wrong view labels is possible, but time-consuming considering the overwhelming number of images involved. ResNet50 model 1 from the study was used to identify potentially abnormal images, from which a total of 83 images with severe quality issues were annotated after human cross-checking. The same principle was similarly applied to correcting mislabelled views using model 2, with 1,054 total corrections. Additionally, model 2 was utilised to classify images previously without view labels with human crosscheck. The augmented image metadata dataset represents an improvement over the original, serving to maximise the exclusion of images of bad quality or incorrect views in datasets used in future studies.

Conversely, the ResNet50 models themselves may still have room for improvement. Judging from model attention heatmaps, model 1 frequently displays a stronger focus around the edge of the image as opposed to inner zones, which may limit its ability to identify certain types of abnormal images. The hierarchical nature of the models makes model 1 a potential weak spot, as its moderate false negative rate increases the chance of an abnormal image continuing through the CAD system unchecked. However, since abnormal images are rare by nature, the overall impact is likely limited.

### Cardiothoracic Segmentation and Cardiomegaly Detection

Our computer vision model, developed as part of the CAD system, exhibited good performance for binary cardiomegaly classification. As it was not a machine learning model and instead relied on statistical methods, the need for curating a training set is eliminated, and all images within the MIMICCXR dataset can potentially serve as part of the test set. It has an overall accuracy of 95.2% (0.95 AUC) by using all images with unequivocal heart size mentions from MIMICCXR as the test set, utilising MIMIC-CXR report diagnosis as ground truth and 0.51 CV-CTR as the threshold for cardiomegaly (borderline cases are excluded as they do not fit into either category). The CAD model has a false negative rate of 6.6%, indicating that it captures the majority of cardiomegaly cases, while its false positive rate is 3.2% is reflective of high precision of cardiomegaly detection. Note that these numbers may not fully reflect the model’s true accuracy, as the model is not yet tested on other datasets. Also, not all cardiomegaly diagnoses from MIMIC-CXR are necessarily correct, as evidenced later. In terms of overall accuracy, our model proves superior to nearly all existing classification-based (28) and segmentation-based (27, 30, 31) models (Table 3). The test set used in this study (n=19,144) also vastly outnumbers those used in similar studies (131 to 600) by magnitudes in terms of total images included.

**Table 3.** Comparison of binary cardiomegaly classification performance between existing models (as reported in publication) and model in the current study. AUC = Area Under (ROC) Curve. * indicates that the value represents the performance of the best-performing model within the study or the performance measured on a specific test set where the model is most accurate. The top result is indicated in bold, second best underlined.

| Study | Test data | Size | AUC | Accuracy (%) |
| --- | --- | --- | --- | --- |
| (28) | ChestX-ray8 | 600 | 0.87 | 79 |
| (31) | Local hospital | 418 | **0.96\*** | 91 |
| (30) | Local hospital | 183 | N/A | <u>94.9</u> |
| (27) | Local hospital | 131 | 0.92* | 92.3 |
| Ours | MIMIC-CXR | 19,144 | <u>0.95</u> | **95.2** |

Furthermore, while these past studies have developed similar CAD systems for binary cardiomegaly detection, few have gone to lengths to implement functions for differentiating between different levels of severity. In contrast, when radiologists report findings of cardiomegaly, it is often standard practice to attach a severity descriptor. The current study is unique in that it identified the most common descriptors used, grouped synonyms based on statistical analysis and derived CV-CTR intervals for each severity category, which makes the CAD output more informative and is indispensable for downstream report sentence generation.

Our work also explores the subjectivity in severity attribution in clinical practice. Without any universal guidelines for descriptor selection, radiologists rely on individual judgment to pick the most appropriate descriptors. This is a process that permits a significant degree of subjectivity, which is reflected by the wide distributions of CV-CTR across different descriptors and severity categories (Fig 4B). While fluctuations in segmentation and computer vision model performance certainly do contribute to the overlaps, natural subjectivity and personal biases of radiologists are also likely factors.

Furthermore, responses gathered from the radiologist questionnaire revealed 4 cases where the responding radiologist rejected the original MIMIC-CXR judgement of cardiomegaly, which constitutes 12.5% of all original cardiomegaly sentences assessed. In nearly half (47.5%) of all cardiomegaly sentences included in the questionnaire, the two assessing radiologists disagreed in their response agreement levels. Indeed, the strict agreement between assessing radiologists is only 0.4878 (0.864 relaxed), indicating a considerable degree of subjectivity in CXR interpretation and reporting. Nonetheless, a point of positivity lies in the fact that the upper threshold of normal CV-CTR determined by statistical analysis (0.51) is nearly identical to the currently used clinical threshold (0.5), indicating a high level of adherence in clinical practice.

In addition, studies have shown that CTR may have limited diagnostic power when compared to cardiac MRI, the gold standard for cardiac chamber enlargement detection (48, 49). Therefore, we have explored the CAR as an alternative metric for cardiomegaly detection. While not used clinically, the concept considers the definition and presentation of cardiomegaly and has the advantage of considering two dimensions of the heart as opposed to a single dimension used for CTR calculation. Despite these advantages, we did not observe a major difference between CTR and CAR in terms of alignment with cardiomegaly severity. However, the reliability of the CAD system may be improved by cross-checking CV-CTR and CV-CAR derived from the same images, flagging instances where a significant mismatch is found between predicted severity by both methods.

The study also examined correlations between cardiac enlargement and various comorbidities. The mention of edema appeared to be the strongest predictor for enlarged cardiac size, with CV-CTR greater than 0.5 in over 90% of cases, only marginally less than mentioning of cardiomegaly itself. As such, the presence edema may be strongly predictive of the co-occurrence of cardiomegaly. This reflected the outcome of the co-occurrence analysis showing a frequent cooccurrence between the two conditions (Fig 6B).

### LLM Sentence Generation and CAD Performance Evaluation

The inclusion of an automatic report sentence generation module is another feature less commonly found in other cardiomegaly CAD systems, the addition of which further enhances user experience by removing the need for manual text input. Compared to more diverse and complex conditions, the reporting of cardiomegaly has the advantage of being quite consistent and uncomplicated, which enables LLMs to imitate their style effectively.

Another strength of the study is the inclusion of radiologist assessment of the performance of the CAD system, a rare inclusion with parallels only found in one other study (30). Whereas most other work in the space of report generation use natural language generation metrics to evaluate the agreement between generated text and original reports (50–52). While the assessment is limited in scale, it nevertheless helped validate the system’s feasibility. The current CAD system reached an average agreement of S:0.545/R:0.929 with our radiologists’ verdicts, comparable with S:0.624/R:0.845 between our radiologists and MIMIC, suggesting that the CAD system’s performance may be non-inferior, or share a comparable level of subjectivity, to trained radiologists. Our findings indicate that our machine-generated sentences were on par with the MIMIC radiologists in assessing the severity of cardiomegaly and the semantics used to generate the report. In fact, disagreements appear to be mostly linguistic rather than diagnostic, as evidenced by the distribution of reasons for disagreement cited, which can be improved by curating a better set of generated sentences.

### Limitations and Future Work

While the preliminary results offer much encouragement, the study remains a proof-of-principle at this stage, owing to the lack of large-scale trials involving clinical validation. It is also yet to be tested on datasets other than MIMIC-CXR. Furthermore, the caseby-case performance of the computer vision model is dependent on accurate anatomical segmentations, and as such is vulnerable to errors arising from aberrant segmentations. It is especially a concern among CXR images where the cardiac silhouette and/or part of the lungs is obscured by conditions such as pulmonary edema and pleural effusion, which can significantly diminish the reliability of segmentations obtained with CXAS. A more versatile segmentation model optimised for challenging CXR images could greatly benefit the performance of the model.

Additionally, since PA is the recommended view for cardiomegaly detection, AP CXR images have been excluded from analysis, despite the close resemblance of AP CXR to PA CXR. While AP CXR has the inclination to exaggerate cardiac size (53), they can, nevertheless, serve as an initial screening for determining the need for further investigation. For instance, one study has recommended a CTR ratio of 0.6 as the threshold when using AP CXR for increased specificity (54). The CAD’s usage scenarios may therefore be expanded by including AP CXR in follow-up studies.

During the preparatory phase in LLM sentence generation, it was noted that a significant portion of cardiomegaly mentions includes references to past time points, examples include ‘mild cardiomegaly is unchanged’ and ‘severe cardiomegaly has improved’. The study did not explore the possibility of generating sentences capturing multiple time points. However, it may be feasible by allowing the model to calculate CTR and make a severity prediction for both the past and present CXR in question, using any difference as an indicator of change to generate more dynamic sentences.

### Conclusion

The present study has developed the essential framework and components for the assembly of a highly automated and reliable CAD system for cardiomegaly detection using PA CXR images. The system holds several advantages, including a high degree of cost-effectiveness, explainability in detection, and efficiency in operation. Input CXR images are first screened to exclude and flag instances unsuitable for cardiomegaly detection. By calculating CTR using a computer vision model from deep learning-derived heart and lung segmentations, high overall accuracy in detecting cardiomegaly and assigning appropriate severity labels is achieved. Furthermore, the model is capable of generating human-like, report-styled sentences based on the output of the computer vision model. Performance metrics indicate that the ability of the model to generate accurate cardiomegaly report sentences based on PA CXR is likely to be non-inferior to human radiologists, although larger trials are necessary to confirm this judgment. With further optimisation, the novel CAD system could prove to be a welcome addition to the radiology toolkit, representing an accessible and cost-effective method to offer radiologists a second perspective to validate their own assessments.

## Data Availability

All data produced in the present work are contained in the manuscript.

## AUTHOR CONTRIBUTIONS

Conceptualisation, J.M.P. and M.G.S.; Methodology, T.Z., K.X., A.D.L. and J.M.P.; Formal Analysis and Investigation, T.Z., K.X. and W.S.; Supervision, J.M.P., A.D.L., M.G.S., K.L.R. and M.B.M.; Writing – Original Draft: T.Z., K.X., A.D.L. and J.M.P.; Writing – Review & Editing: J.M.P., A.D.L., K.L.R., and M.G.S. All authors have read and agreed to the published version of the manuscript.

## Acknowledgement

We gratefully acknowledge the support of Dr Catherine Payne, Dr Abi Redwood, Dr Liam Roebuck and Dr John Taylor (listed alphabetically) in scoring the reports. We would also like to express our gratitude to Dr Jamie Campbell and Dr Joanna Davis for their time and assistance in gathering expert feedback. We thank Dr Tom Pollard and Dr Alistair Johnson for their advice and clarifications on the MIMIC-CXR data user agreement that prohibits disclosure of data (images, report snippets). The NIH Clinical Center was the provider of the de-identified images of chest xrays shown in this manuscript, and is available through the NIH download site: https://nihcc.app.box.com/v/ChestXray-NIHCC.

## Supplementary Materials

**Fig. 7.**
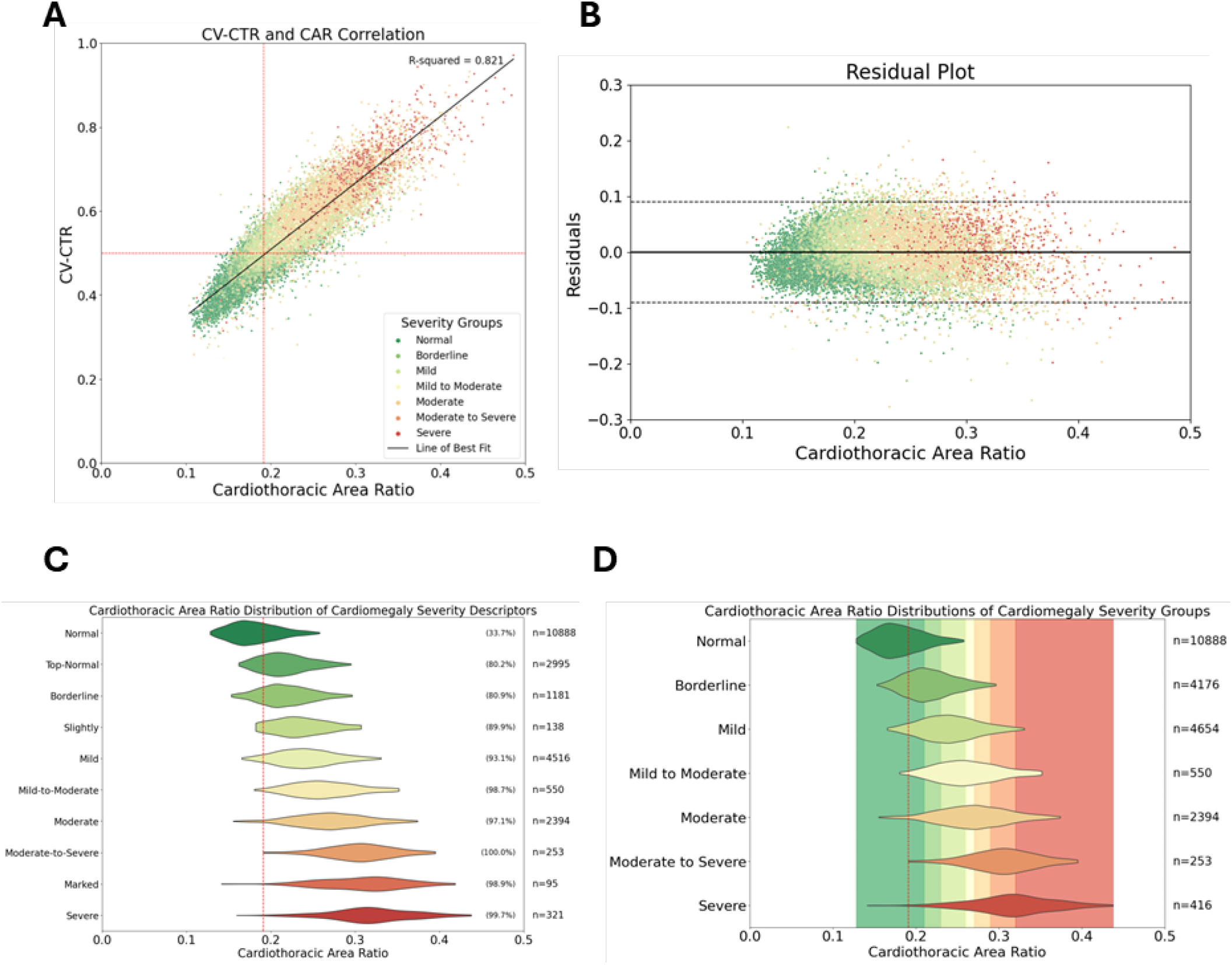
Data-driven severity classification based on natural language extracted labels and computer vision-based cardiothoracic area ratio (CAR) calculation. A: CV-CAR vs CV-CTR with the 7 severity groups from reports indicated by colours. The horizontal red dashed line represents CV-CTR=0.50 (common threshold for cardiomegaly), the vertical red dashed line represents the CAR value for the lowest 1% of all cardiomegaly cases (from mild to severe), with 99% of cardiomegaly cases to be found above this threshold. B: Residual plot based on the line of best fit. The parallel black dashed lines represents the interval where 90% of residuals reside. C: Violin plot of the CV-CAR for all categories prior to merging categories. D: Violin plot of the CV-CAR after consolidation of categories. C,D: The red dashed line represents the 1% CAR value threshold for cardiomegaly cases (from mild to severe, as in panel A).

## Bibliography

1. Anouk M Speets, Yolanda van der Graaf, Arno W Hoes, Sandra Kalmijn, Alfred PE Sachs, Matthieu JCM Rutten, Jan Willem C Gratama, Alexander D Montauban van Swijndregt, and Willem PThM Mali. Chest radiography in general practice: indications, diagnostic yield and consequences for patient management. British Journal of General Practice, 56(529): 574–578, 2006. ISSN 0960-1643.

2. S. Skinner. Guide to thoracic imaging. Aust Fam Physician, 44(8):558–62, 2015.

3. Cornelia Schaefer-Prokop, Ulrich Neitzel, Henk W. Venema, Martin Uffmann, and Mathias Prokop. Digital chest radiography: an update on modern technology, dose containment and control of image quality. European Radiology, 18(9):1818–1830, April 2008. ISSN 1432-1084. doi: 10.1007/s00330-008-0948-3.

4. H G Chotas and C E Ravin. Chest radiography: estimated lung volume and projected area obscured by the heart, mediastinum, and diaphragm. Radiology, 193(2):403–404, November 1994. ISSN 1527-1315. doi: 10.1148/radiology.193.2.7972752.

5. Catherine M Jones, Quinlan D Buchlak, Luke Oakden-Rayner, Michael Milne, Jarrel Seah, Nazanin Esmaili, and Ben Hachey. Chest radiographs and machine learning – past, present and future. Journal of Medical Imaging and Radiation Oncology, 65(5):538–544, June 2021. ISSN 1754-9485. doi: 10.1111/1754-9485.13274.

6. M.M. Boubnovski, M. Chen, K. Linton-Reid, J.M. Posma, S.J. Copley, and E.O. Aboagye. Development of a multi-task learning v-net for pulmonary lobar segmentation on ct and application to diseased lungs. Clinical Radiology, 77(8):e620–e627, August 2022. ISSN 0009-9260. doi: 10.1016/j.crad.2022.04.012.

7. Stephen Waite, Jinel Scott, Brian Gale, Travis Fuchs, Srinivas Kolla, and Deborah Reede. Interpretive error in radiology. American Journal of Roentgenology, 208(4):739–749, April 2017. ISSN 1546-3141. doi: 10.2214/ajr.16.16963.

8. Andrew J. Degnan, Emily H. Ghobadi, Peter Hardy, Elizabeth Krupinski, Elena P. Scali, Lindsay Stratchko, Adam Ulano, Eric Walker, Ashish P. Wasnik, and William F. Auffermann. Perceptual and interpretive error in diagnostic radiology—causes and potential solutions. Academic Radiology, 26(6):833–845, June 2019. ISSN 1076-6332. doi: 10.1016/j.acra.2018.11.006.

9. V. Markotic, T. Pojuzina, D. Radancevic, M. Miljko, and V. Pokrajcic. The radiologist workload increase; where is the limit?: Mini review and case study. Psychiatr Danub, 33(Suppl 4): 768–770, 2021.

10. Jay R. Parikh, Darcy Wolfman, Claire E. Bender, and Elizabeth Arleo. Radiologist burnout according to surveyed radiology practice leaders. Journal of the American College of Radiology, 17(1):78–81, January 2020. ISSN 1546-1440. doi: 10.1016/j.jacr.2019.07.008.

11. Efosa P. Iyawe, Bukunmi M. Idowu, and Olasubomi J. Omoleye. Radiology subspecialisation in africa: A review of the current status. South African Journal of Radiology, 25(1), August 2021. ISSN 1027-202X. doi: 10.4102/sajr.v25i1.2168.

12. Adrian P. Brady. Error and discrepancy in radiology: inevitable or avoidable? Insights Into Imaging, 8(1):171–182, December 2016. ISSN 1869-4101. doi: 10.1007/s13244-016-0534-1.

13. Maya Pavlova, Naomi Terhljan, Audrey G. Chung, Andy Zhao, Siddharth Surana, Hossein Aboutalebi, Hayden Gunraj, Ali Sabri, Amer Alaref, and Alexander Wong. Covid-net cxr-2: An enhanced deep convolutional neural network design for detection of covid-19 cases from chest x-ray images. Frontiers in Medicine, 9, June 2022. ISSN 2296-858X. doi: 10.3389/fmed.2022.861680.

14. Ajay Sharma and Pramod Kumar Mishra. Covid-manet: Multi-task attention network for explainable diagnosis and severity assessment of covid-19 from cxr images. Pattern Recognition, 131:108826, November 2022. ISSN 0031-3203. doi: 10.1016/j.patcog.2022.108826.

15. Bram van Ginneken. Fifty years of computer analysis in chest imaging: rule-based, machine learning, deep learning. Radiological Physics and Technology, 10(1):23–32, February 2017. ISSN 1865-0341. doi: 10.1007/s12194-017-0394-5.

16. Jun Gao, Qian Jiang, Bo Zhou, and Daozheng Chen. Convolutional neural networks for computer-aided detection or diagnosis in medical image analysis: An overview. Mathematical Biosciences and Engineering, 16(6):6536–6561, 2019. ISSN 1551-0018. doi: 10.3934/mbe.2019326.

17. Yan-Wei Lee, Sheng-Kai Huang, and Ruey-Feng Chang. Chexgat: A disease correlation-aware network for thorax disease diagnosis from chest x-ray images. Artificial Intelligence in Medicine, 132:102382, October 2022. ISSN 0933-3657. doi: 10.1016/j.artmed.2022.102382.

18. Saleh Albahli and Tahira Nazir. Ai-centernet cxr: An artificial intelligence (ai) enabled system for localization and classification of chest x-ray disease. Frontiers in Medicine, 9, August 2022. ISSN 2296-858X. doi: 10.3389/fmed.2022.955765.

19. Marc Boubnovski Martell, Kristofer Linton-Reid, Sumeet Hindocha, Mitchell Chen, Paula Moreno, Marina Álvarez-Benito, Ángel Salvatierra, Richard Lee, Joram M. Posma, Marco A. Calzado, and Eric O. Aboagye. Deep representation learning of tissue metabolome and computed tomography annotates nsclc classification and prognosis. npj Precision Oncology, 8(1), February 2024. ISSN 2397-768X. doi: 10.1038/s41698-024-00502-3.

20. Hina Amin and Waqas J. Siddiqui. Cardiomegaly. StatPearls Publishing, 2024.

21. John Lynn Jefferies and Jeffrey A Towbin. Dilated cardiomyopathy. The Lancet, 375(9716): 752–762, February 2010. ISSN 0140-6736. doi: 10.1016/s0140-6736(09)62023-7.

22. Michinari Nakamura and Junichi Sadoshima. Mechanisms of physiological and pathological cardiac hypertrophy. Nature Reviews Cardiology, 15(7):387–407, April 2018. ISSN 1759-5010. doi: 10.1038/s41569-018-0007-y.

23. Ippei Shimizu and Tohru Minamino. Physiological and pathological cardiac hypertrophy. Journal of Molecular and Cellular Cardiology, 97:245–262, August 2016. ISSN 0022-2828. doi: 10.1016/j.yjmcc.2016.06.001.

24. G. Michael Felker, Richard E. Thompson, Joshua M. Hare, Ralph H. Hruban, Diedre E. Clemetson, David L. Howard, Kenneth L. Baughman, and Edward K. Kasper. Underlying causes and long-term survival in patients with initially unexplained cardiomyopathy. New England Journal of Medicine, 342(15):1077–1084, April 2000. ISSN 1533-4406. doi: 10.1056/nejm200004133421502.

25. Rohit S Loomba, Parinda H Shah, Karan Nijhawan, Saurabh Aggarwal, and Rohit Arora. Cardiothoracic ratio for prediction of left ventricular dilation: A systematic review and pooled analysis. Future Cardiology, 11(2):171–175, March 2015. ISSN 1744-8298. doi: 10.2217/fca.15.5.

26. Krystian Truszkiewicz, Rafał Poręba, and Paweł Gać. Radiological cardiothoracic ratio in evidence-based medicine. Journal of Clinical Medicine, 10(9):2016, May 2021. ISSN 2077-0383. doi: 10.3390/jcm10092016.

27. Patrick Thiam, Christopher Kloth, Daniel Blaich, Andreas Liebold, Meinrad Beer, and Hans A. Kestler. Segmentation-based cardiomegaly detection based on semi-supervised estimation of cardiothoracic ratio. Scientific Reports, 14(1), March 2024. ISSN 2045-2322. doi: 10.1038/s41598-024-56079-1.

28. S. Zhou, X. Zhang, and R. Zhang. Identifying cardiomegaly in chestx-ray8 using transfer learning. Stud Health Technol Inform, 264:482–486, 2019. ISSN 1879-8365 (Electronic), 0926-9630 (Linking). doi: 10.3233/SHTI190268.

29. Haralabos Bougias, Eleni Georgiadou, Christina Malamateniou, and Nikolaos Stogiannos. Identifying cardiomegaly in chest x-rays: a cross-sectional study of evaluation and comparison between different transfer learning methods. Acta Radiologica, 62(12):1601–1609, November 2020. ISSN 1600-0455. doi: 10.1177/0284185120973630.

30. Pranav Ajmera, Amit Kharat, Tanveer Gupte, Richa Pant, Viraj Kulkarni, Vinay Duddalwar, and Purnachandra Lamghare. Observer performance evaluation of the feasibility of a deep learning model to detect cardiomegaly on chest radiographs. Acta Radiologica Open, 11 (7):205846012211073, July 2022. ISSN 2058-4601. doi: 10.1177/20584601221107345.

31. Mu Sook Lee, Yong Soo Kim, Minki Kim, Muhammad Usman, Shi Sub Byon, Sung Hyun Kim, Byoung Il Lee, and Byoung-Dai Lee. Evaluation of the feasibility of explainable computer-aided detection of cardiomegaly on chest radiographs using deep learning. Scientific Reports, 11(1), August 2021. ISSN 2045-2322. doi: 10.1038/s41598-021-96433-1.

32. Abbas Jafar, Muhammad Talha Hameed, Nadeem Akram, Umer Waqas, Hyung Seok Kim, and Rizwan Ali Naqvi. Cardionet: Automatic semantic segmentation to calculate the cardiothoracic ratio for cardiomegaly and other chest diseases. Journal of Personalized Medicine, 12(6):988, June 2022. ISSN 2075-4426. doi: 10.3390/jpm12060988.

33. Muhammad Arsalan, Muhammad Owais, Tahir Mahmood, Jiho Choi, and Kang Ryoung Park. Artificial intelligence-based diagnosis of cardiac and related diseases. Journal of Clinical Medicine, 9(3):871, March 2020. ISSN 2077-0383. doi: 10.3390/jcm9030871.

34. Xiaosong Wang, Yifan Peng, L. Lu, Zhiyong Lu, Mohammadhadi Bagheri, and Ronald M. Summers. Chestx-ray8: Hospital-scale chest x-ray database and benchmarks on weakly-supervised classification and localization of common thorax diseases. In 2017 IEEE Conference on Computer Vision and Pattern Recognition (CVPR). IEEE, July 2017. doi: 10.1109/cvpr.2017.369.

35. Dong Hwan Kim, Seung Don Yoo, Sung Min Kim, Sung Jig Im, and Dong Whan Hong. Atypical thoracic solitary plasmacytoma. Annals of Rehabilitation Medicine, 36(5):739, 2012. ISSN 2234-0653. doi: 10.5535/arm.2012.36.5.739.

36. Alistair E. W. Johnson, Tom J. Pollard, Seth J. Berkowitz, Nathaniel R. Greenbaum, Matthew P. Lungren, Chih-ying Deng, Roger G. Mark, and Steven Horng. Mimic-cxr, a deidentified publicly available database of chest radiographs with free-text reports. Scientific Data, 6(1), December 2019. ISSN 2052-4463. doi: 10.1038/s41597-019-0322-0.

37. A. L. Goldberger, L. A. N. Amaral, L. Glass, J. M. Hausdorff, P. Ch. Ivanov, R. G. Mark, J. E. Mietus, G. B. Moody, C.-K. Peng, and H. E. Stanley. Physiobank, physiotoolkit, and physionet: Components of a new research resource for complex physiologic signals. Circulation, 101(23):e215–e220, June 2000. doi: 10.1161/01.CIR.101.23.e215.

38. Jeremy Irvin, Pranav Rajpurkar, Michael Ko, Yifan Yu, Silviana Ciurea-Ilcus, Chris Chute, Henrik Marklund, Behzad Haghgoo, Robyn Ball, Katie Shpanskaya, Jayne Seekins, David A. Mong, Safwan S. Halabi, Jesse K. Sandberg, Ricky Jones, David B. Larson, Curtis P. Langlotz, Bhavik N. Patel, Matthew P. Lungren, and Andrew Y. Ng. Chexpert: A large chest radiograph dataset with uncertainty labels and expert comparison. Proceedings of the AAAI Conference on Artificial Intelligence, 33(01):590–597, July 2019. ISSN 2159-5399. doi: 10.1609/aaai.v33i01.3301590.

39. Yifan Peng, Xiaosong Wang, L. Lu, Mohammadhadi Bagheri, Ronald M. Summers, and Zhiyong Lu. Negbio: a high-performance tool for negation and uncertainty detection in radiology reports. AMIA Summits on Translational Science Proceedings, pages 188–196, May 2018.

40. Kaiming He, Xiangyu Zhang, Shaoqing Ren, and Jian Sun. Deep residual learning for image recognition. arXiv, December 2015. doi: 10.48550/ARXIV.1512.03385.

41. Ramprasaath R. Selvaraju, Michael Cogswell, Abhishek Das, Ramakrishna Vedantam, Devi Parikh, and Dhruv Batra. Grad-cam: Visual explanations from deep networks via gradient-based localization. In 2017 IEEE International Conference on Computer Vision (ICCV), pages 618–626, October 2017. doi: 10.1109/ICCV.2017.74.

42. Constantin Seibold, Alexander Jaus, Matthias A. Fink, Moon Kim, Simon Reiß, Ken Herrmann, Jens Kleesiek, and Rainer Stiefelhagen. Accurate fine-grained segmentation of human anatomy in radiographs via volumetric pseudo-labeling. arXiv, 2023. doi: 10.48550/ARXIV.2306.03934.

43. Tao Huang, Rui Yang, Longbin Shen, Aozi Feng, Li Li, Ningxia He, Shuna Li, Liying Huang, and Jun Lyu. Deep transfer learning to quantify pleural effusion severity in chest x-rays. BMC Medical Imaging, 22(1), May 2022. ISSN 1471-2342. doi: 10.1186/s12880-022-00827-0.

44. Wilson Silva, Tiago Gonçalves, Kirsi Härmä, Erich Schröder, Verena Carola Obmann, María Cecilia Barroso, Alexander Poellinger, Mauricio Reyes, and Jaime S. Cardoso. Computer-aided diagnosis through medical image retrieval in radiology. Scientific Reports, 12(1), December 2022. ISSN 2045-2322. doi: 10.1038/s41598-022-25027-2.

45. Judy Wawira Gichoya, Imon Banerjee, Ananth Reddy Bhimireddy, John L Burns, Leo Anthony Celi, Li-Ching Chen, Ramon Correa, Natalie Dullerud, Marzyeh Ghassemi, Shih-Cheng Huang, Po-Chih Kuo, Matthew P Lungren, Lyle J Palmer, Brandon J Price, Saptarshi Purkayastha, Ayis T Pyrros, Lauren Oakden-Rayner, Chima Okechukwu, Laleh Seyyed-Kalantari, Hari Trivedi, Ryan Wang, Zachary Zaiman, and Haoran Zhang. Ai recognition of patient race in medical imaging: a modelling study. The Lancet Digital Health, 4(6): e406–e414, June 2022. ISSN 2589-7500. doi: 10.1016/s2589-7500(22)00063-2.

46. Zhihong Lin, Donghao Zhang, Danli Shi, Renjing Xu, Qingyi Tao, Lin Wu, Mingguang He, and Zongyuan Ge. Contrastive pre-training and linear interaction attention-based transformer for universal medical reports generation. Journal of Biomedical Informatics, 138: 104281, February 2023. ISSN 1532-0464. doi: 10.1016/j.jbi.2023.104281.

47. Qingqing Zhu, Tejas Sudharshan Mathai, Pritam Mukherjee, Yifan Peng, Ronald M. Summers, and Zhiyong Lu. Utilizing longitudinal chest x-rays and reports to pre-fill radiology reports. arXiv, 2023. doi: 10.48550/ARXIV.2306.08749.

48. Reghunath Anjuna, Simkus Paulius, Gutierrez Gimeno Manuel, Banisauskaite Audra, Noreikaite Jurate, and Radike Monika. Diagnostic value of cardiothoracic ratio in patients with non-ischaemic cardiomyopathy: comparison to cardiovascular magnetic resonance imaging. Current Problems in Diagnostic Radiology, 53(3):353–358, May 2024. ISSN 0363-0188. doi: 10.1067/j.cpradiol.2024.01.011.

49. Paulius Simkus, Manuel Gutierrez Gimeno, Audra Banisauskaite, Jurate Noreikaite, David McCreavy, Diana Penha, and Monika Arzanauskaite. Limitations of cardiothoracic ratio derived from chest radiographs to predict real heart size: comparison with magnetic resonance imaging. Insights into Imaging, 12(1), November 2021. ISSN 1869-4101. doi: 10.1186/s13244-021-01097-0.

50. Tim Tanida, Philip Müller, Georgios Kaissis, and Daniel Rueckert. Interactive and explainable region-guided radiology report generation. In 2023 IEEE/CVF Conference on Computer Vision and Pattern Recognition (CVPR). IEEE, June 2023. doi: 10.1109/cvpr52729.2023.00718.

51. Aaron Nicolson, Jason Dowling, and Bevan Koopman. Improving chest x-ray report generation by leveraging warm starting. Artificial Intelligence in Medicine, 144:102633, October 2023. ISSN 0933-3657. doi: 10.1016/j.artmed.2023.102633.

52. Jean-Benoit Delbrouck, Maya Varma, Pierre Chambon, and Curtis Langlotz. Overview of the radsum23 shared task on multi-modal and multi-anatomical radiology report summarization. In The 22nd Workshop on Biomedical Natural Language Processing and BioNLP Shared Tasks. Association for Computational Linguistics, 2023. doi: 10.18653/v1/2023.bionlp-1.45.

53. M.T. Tsakok and F.V. Gleeson. The chest radiograph in heart disease. Medicine, 46(8): 453–457, August 2018. ISSN 1357-3039. doi: 10.1016/j.mpmed.2018.05.007.

54. Hakan Sahin, Divya N. Chowdhry, Andrew Olsen, Omar Nemer, and Lindsay Wahl. Is there any diagnostic value of anteroposterior chest radiography in predicting cardiac chamber enlargement? The International Journal of Cardiovascular Imaging, 35(1):195–206, August 2018. ISSN 1573-0743. doi: 10.1007/s10554-018-1442-x.

